# Virtual Spectral Decomposition with Dendritic Binary Gating Detects Pancreatic Cancer Tissue Transformation on Standard CT: Multi-Institutional Validation Across Three Independent Datasets with Evidence of Temporally Stable Pre-Pathological Signal

**DOI:** 10.64898/2026.04.08.26350418

**Authors:** Shubham Chandra

## Abstract

**Background:** Pancreatic ductal adenocarcinoma (PDAC) has a five-year survival rate of approximately 12%, largely because it is typically diagnosed at an advanced stage. CT-based computational methods for early detection exist but rely on black-box deep learning or large texture feature sets without tissue-specific interpretability. A method that decomposes standard CT into named tissue-component channels with full explainability would bridge the gap between computational detection and clinical understanding.

**Methods:** We developed Virtual Spectral Decomposition (VSD), which applies six parameterized sigmoid functions S(HU) = 1/(1+exp(−α(HU−μ))) to standard portal-venous CT, decomposing each pixel into tissue-specific response channels for fat (μ=−60), fluid (μ=10), parenchyma (μ=45), stroma (μ=75), vascular (μ=130), and calcification (μ=250). Dendritic Binary Gating identifies structural content per channel using morphological filtering, enabling co-firing analysis and lone firer identification. A 25-feature signature was extracted per patient. Three independent datasets were analyzed: NIH Pancreas-CT (n=78 healthy), Medical Segmentation Decathlon Task07 (n=281 PDAC, paired tumor/adjacent tissue), and CPTAC-PDA from The Cancer Imaging Archive (n=82, multi-institutional, with DICOM time point tags encoding days relative to pathological diagnosis). The same six sigmoid parameters were used across all datasets without retraining.

**Results:** VSD achieved AUC 0.943 for field effect detection (healthy vs. cancer-adjacent parenchyma) and AUC 0.931 for patient-stratified tumor specification on MSD. On CPTAC-PDA, VSD achieved AUC 0.961 (6 features) and 0.979 (25 features) for distinguishing healthy from cancer-bearing pancreas. All significant features replicated across datasets in the same direction: z_fat (d=−2.10, p=3.5×10^−27^), z_fluid (d=−2.76, p=2.4×10^−38^), fire_fat (d=+2.18, p=1.2×10^−28^). VSD severity showed no correlation with days-from-pathological-diagnosis (r=−0.008, p=0.944) across a range of day −1,394 to day +249, indicating a temporally stable tissue state. In an exploratory observation, patient C3N-01375, scanned 1,394 days (3.8 years) before pathological confirmation, showed VSD severity 2.8 standard deviations above the healthy mean.

**Conclusions:** VSD with Dendritic Binary Gating detects a stable pancreatic tissue composition signature on standard CT that is present on pre-pathological imaging, validated across three independent datasets without parameter adjustment. The six sigmoid channels map to biologically meaningful tissue components through a fully transparent interpretability chain. The temporal stability of the signal across available time points suggests that VSD detects an early, persistent tissue state rather than a progressively worsening process. If confirmed in prospective cohorts with truly incidental pre-diagnostic CTs, VSD could function as a single-scan screening tool applicable to abdominal CT performed during the pre-clinical window. Prospective longitudinal validation is the critical next step.

## 1. INTRODUCTION

Pancreatic ductal adenocarcinoma (PDAC) remains one of the most lethal malignancies, with a five-year survival rate of approximately 12% and projected to become the second leading cause of cancer death in the United States by 2030. The dismal prognosis is driven by late-stage diagnosis: over 80% of patients present with locally advanced or metastatic disease because early PDAC produces no symptoms and no detectable mass on standard imaging.

The biological window for detection, however, is surprisingly long. PDAC develops through a well-characterized precursor progression from PanIN-1 through PanIN-3 (carcinoma in situ) to invasive carcinoma, spanning an estimated 10–15 years. During the PanIN-3 stage, the pancreatic microenvironment undergoes significant remodeling: stromal desmoplasia, acinar atrophy, fatty infiltration, and inflammatory changes alter the tissue composition years before a visible tumor mass forms. These microenvironment changes, while insufficient to produce a mass lesion detectable by the radiologist’s eye, do alter the Hounsfield Unit (HU) distribution of the tissue.

Recent work has demonstrated the feasibility of detecting pre-cancerous pancreatic changes on CT. Cao et al. developed the PANDA deep learning model achieving AUC 0.986 for PDAC detection using large-scale non-contrast CT datasets [7]. Kenner et al. developed the REDMOD radiomics pipeline achieving AUC 0.98 on 88 texture features from patients scanned up to 36 months before diagnosis [8]. Qureshi et al. at Cedars-Sinai demonstrated 86% classification accuracy on CT scans obtained 6 months to 3 years before diagnosis using radiomic texture features in 36 patients [9]. Ahmed et al. reported that pancreatic abnormalities could be retrospectively identified on CT in up to 38% of studies performed 5 years before clinical PDAC diagnosis [10]. However, existing AI approaches share a critical limitation: they operate as black boxes, providing no tissue-specific interpretation of what compositional changes they detect. The 88 REDMOD features are mathematical texture descriptors (GLCM, GLRLM, shape statistics) with no direct mapping to histological tissue components.

We propose Virtual Spectral Decomposition (VSD), a method that exploits the Hounsfield Unit scale as a spectral axis. Just as optical spectroscopy decomposes light into wavelength-specific components corresponding to different chromophores, VSD decomposes the HU axis into tissue-specific components using parameterized sigmoid functions centered at known tissue densities. Each of six sigmoid channels corresponds to a named tissue type: fat, fluid, parenchyma, stroma, vascular, and calcification. Combined with Dendritic Binary Gating—a morphological method that identifies structural content within each tissue channel—VSD produces a 25-feature tissue-composition signature with full explainability: every feature traces from CT pixel through HU value through sigmoid response through tissue identity through clinical finding.

Here we present multi-institutional validation of VSD across three independent datasets, demonstrate temporal stability of the VSD signature on pre-pathological imaging, and establish the foundation for prospective investigation of VSD as a single-scan screening tool.

## 2. MATERIALS AND METHODS

### 2.1 Datasets

**Population A (Healthy Controls): NIH Pancreas-CT.** 78 contrast-enhanced portal-venous abdominal CT volumes from patients without pancreatic pathology, acquired at the NIH Clinical Center. Expert manual segmentation labels provided [4].

**Population B and C (PDAC Patients): Medical Segmentation Decathlon Task07.** 281 portal-venous CT volumes from patients with confirmed PDAC (Memorial Sloan Kettering Cancer Center). Manual segmentation provides voxel-level labels: label=1 (pancreatic parenchyma adjacent to tumor, Population B) and label=2 (tumor, Population C). Populations B and C are paired within patients [5].

**CPTAC-PDA (Multi-Institutional Validation):** 166 histologically confirmed PDAC patients from the Clinical Proteomic Tumor Analysis Consortium, available through The Cancer Imaging Archive [6]. Multi-institutional (University of Calgary and additional sites), heterogeneous scanners and protocols. DICOM tag (0012,0050) encodes days from pathological diagnosis. 82 patients with successful auto-segmentation (TotalSegmentator) were analyzed. *Important caveat:* the CPTAC time point tags measure days relative to pathological confirmation, not days relative to initial clinical suspicion. The majority of scans in this dataset are staging CTs obtained after cancer was clinically suspected but before pathology was finalized. This distinction is critical for interpreting temporal analyses (see Discussion).

### 2.2 Virtual Spectral Decomposition

Each CT pixel with Hounsfield Unit value HU is passed through six parameterized sigmoid functions:

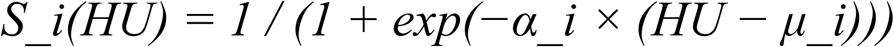

where μ_i is the center of the i-th tissue band and α_i controls the selectivity. The six channels and their parameters are: ch_fat (μ=−60, α=0.05), ch_fluid (μ=10, α=0.04), ch_parenchyma (μ=45, α=0.06), ch_stroma (μ=75, α=0.05), ch_vascular (μ=130, α=0.04), and ch_calcific (μ=250, α=0.02). Parameters were selected based on established HU ranges for each tissue type and held constant across all datasets.

### 2.3 Dendritic Binary Gating

For each VSD channel, structural content is identified using black-tophat morphological filtering (disk structuring element, radius=3 pixels), calibrated against the black-tophat response in reference tissue (non-pancreatic abdominal tissue in the same slice). A pixel is classified as “fired” if its black-tophat response exceeds the reference mean + 0.5 standard deviations. The binary firing map is smoothed with morphological opening and closing (disk, radius=1).

### 2.4 Feature Extraction

Twenty-five features were extracted per patient: (1) six VSD z-scores (mean channel response in pancreas normalized to reference tissue), (2) six Hessian structureness values (multi-scale second-order structure per channel), (3) six firing densities (fraction of pancreatic pixels with structural content per channel), (4) four cross-channel metrics (spectral coherence, lone firer ratio, spectral entropy, binary agreement), and (5) three tissue ratios (fat/stroma, parenchyma/stroma, stroma/vascular). For pixel-level classification, the same 25 features were computed per pixel using local reference normalization.

### 2.5 Auto-Segmentation for CPTAC

CPTAC DICOM files were converted to NIfTI using dcm2niix. Pancreas segmentation was performed using TotalSegmentator (fast mode, pancreas ROI only). Series selection prioritized portal-venous axial acquisitions with ≥60 slices based on series description keyword scoring.

### 2.6 Statistical Analysis

Classification was performed using Random Forest (200 trees, max depth=12) with 5-fold stratified cross-validation and pixel-level evaluation. Effect sizes are reported as Cohen’s d. Group comparisons used Welch’s t-test. Temporal correlations used Spearman rank correlation. Patient-stratified analysis (B vs. C) used GroupKFold to prevent within-patient leakage. VSD severity was defined as the root-mean-square z-score distance from the healthy reference population across all 25 features.

## 3. RESULTS

### 3.1 Field Effect Detection (NIH vs. MSD)

VSD distinguished healthy pancreas (Population A, n=78) from cancer-adjacent parenchyma (Population B, n=281) with AUC 0.943 using all 25 features and AUC 0.926 using only the 6 VSD z-scores (Figure 2). The dominant discriminative features were z_fat (importance 23.9%), z_fluid (15.4%), z_parenchyma (11.7%), structural fat (9.4%), and z_calcific (7.7%).

**Figure 1.**
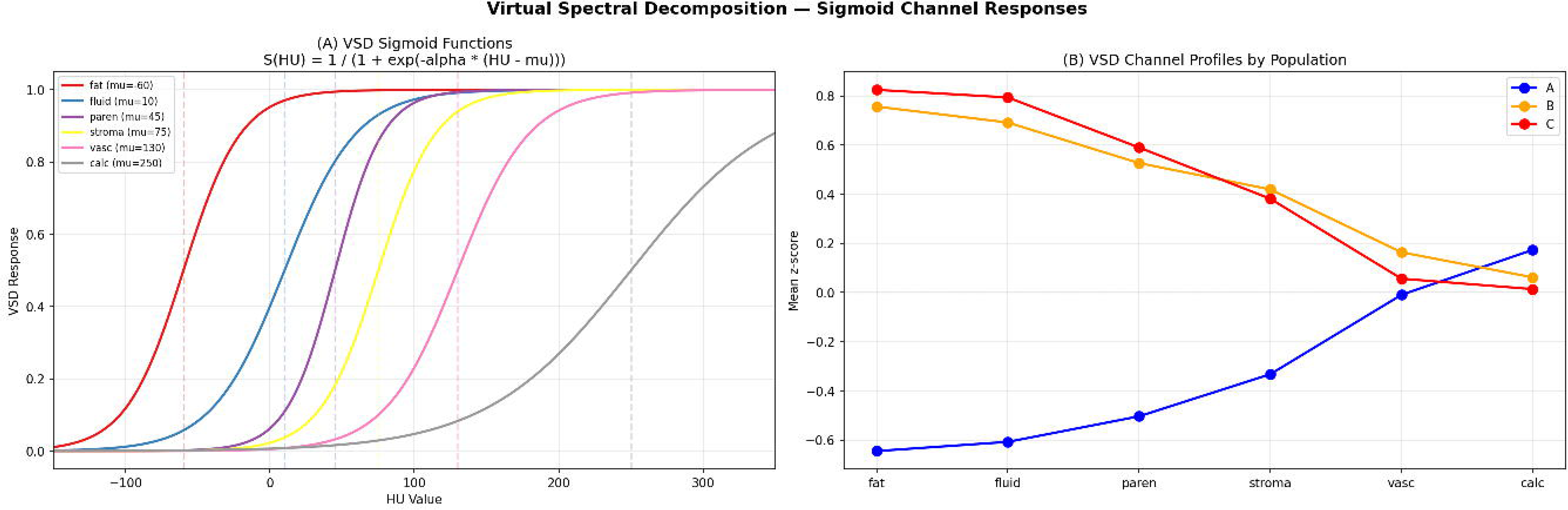
VSD Sigmoid Response Functions and Channel Profiles. (A) Six sigmoid curves showing tissue-specific response across the HU spectrum. (B) Mean VSD z-scores per population demonstrating separation between healthy (blue) and cancer (red) tissue in the fat, fluid, parenchyma, and stroma channels.

**Figure 2.**
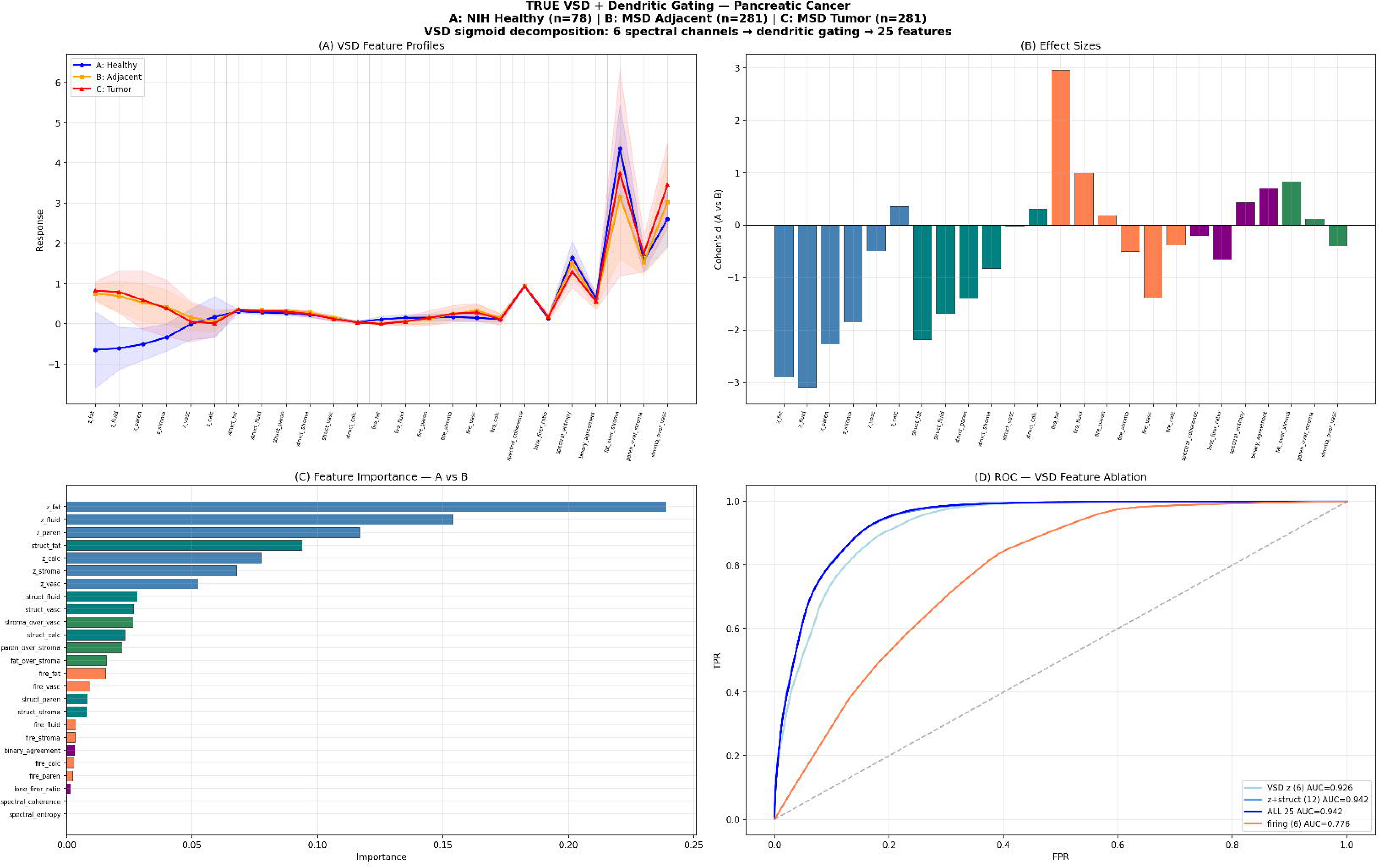
VSD + Dendritic Main Results on MSD Task07. (A) Feature profiles across populations A (healthy), B (adjacent), and C (tumor). (B) Effect sizes (Cohen’s d) across all 25 features for A-vs-B comparison. (C) Feature importance—z_fat dominates at 24%. (D) ROC curves for A-vs-B (AUC 0.943) and B-vs-C (AUC 0.931), with feature ablation showing VSD z-scores alone achieve AUC 0.926.

### 3.2 Patient-Stratified Tumor Specification (MSD)

Using GroupKFold cross-validation with patient ID as the group variable (preventing any within-patient leakage), VSD distinguished adjacent parenchyma (B) from tumor tissue (C) within the same patients with AUC 0.931. The top discriminative features shifted to spectral_entropy (22.1%), structural_fat (14.3%), and firing_stroma (10.2%)—structural and firing features rather than composition z-scores, indicating that B-vs-C discrimination requires architectural analysis beyond tissue composition.

### 3.3 Lone Firer Signature and Structural Coherence

Lone firer analysis revealed a characteristic tissue-component fingerprint of malignant transformation (Figure 3). In healthy pancreas, fat dominated lone-fired pixels at 36%, reflecting the interlobular fat architecture that defines normal pancreatic texture. In cancer-adjacent tissue, vascular lone firing surged to 69%, indicating isolated neovascularization without surrounding organized architecture. In tumor tissue, vascular (45%) and stroma (32%) together accounted for 77% of lone firers, consistent with desmoplastic stroma and tumor vasculature as the dominant surviving structural features.

**Figure 3.**
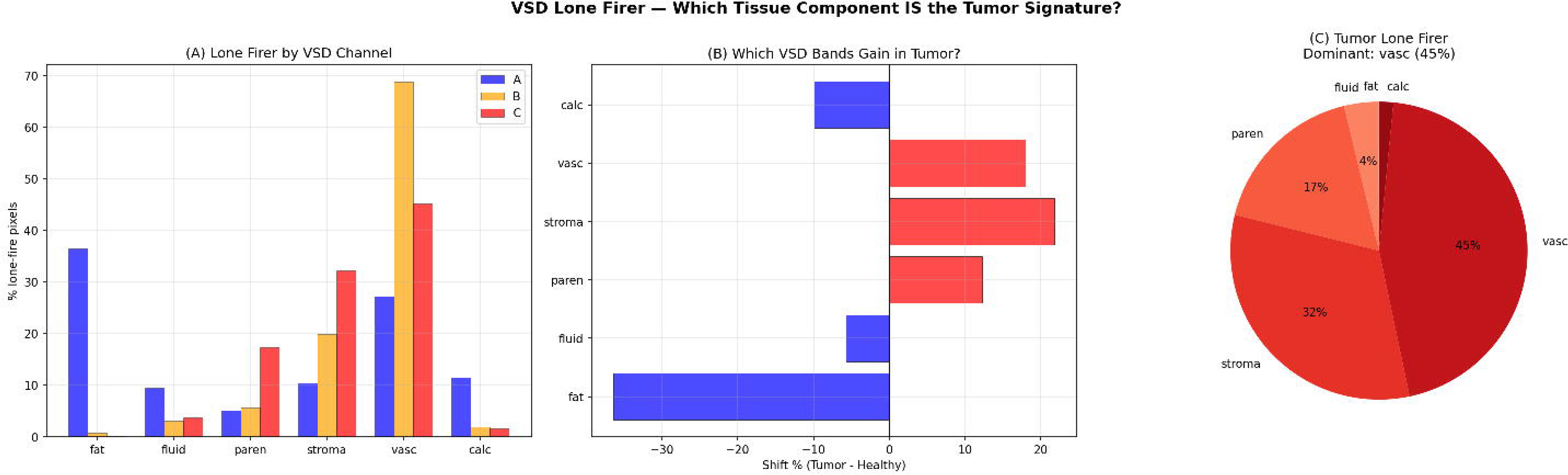
Lone Firer Analysis. (A) Distribution of which VSD channel dominates isolated structural firing across populations. Healthy: fat-dominant (36%). Adjacent: vascular-dominant (69%). Tumor: vascular (45%) + stroma (32%). (B) Shift in band dominance from healthy to tumor. (C) Tumor lone firer composition pie chart.

Multi-channel co-firing (structural pixels where 2+ VSD channels simultaneously detect content) decreased from 56% in healthy tissue to 26% in tumor, quantifying the collapse of organized multi-component architecture. The dominant healthy co-firing pair was HU+structureness (27.5%), representing real anatomy; the dominant tumor pair was DoGc+coherence (24.3%), representing residual texture without cellular organization.

### 3.4 Four-Stage Progressive Detection Sequence

Progressive detection analysis, which ranked cancer patients from mildest to most severe field effect and identified which features cross statistical significance first, revealed a four-stage transformation sequence (Figure 4). Stage 1 (detectable in the mildest 5%): z_fat, z_fluid, z_parenchyma, z_stroma—composition shift before structural reorganization. Stage 2 (7–10%): struct_fat, struct_fluid, fire_fat—structural remodeling and fat architecture destruction (Figure 5). Stage 3 (10–25%): fire_stroma, fire_vasc, binary_agreement—firing pattern shift with desmoplastic structure formation. Stage 4 (65%+): struct_vasc, spectral_entropy, z_vasc—global tissue reorganization. This sequence is consistent with the known biology of PanIN progression, where composition changes precede structural remodeling.

**Figure 4.**
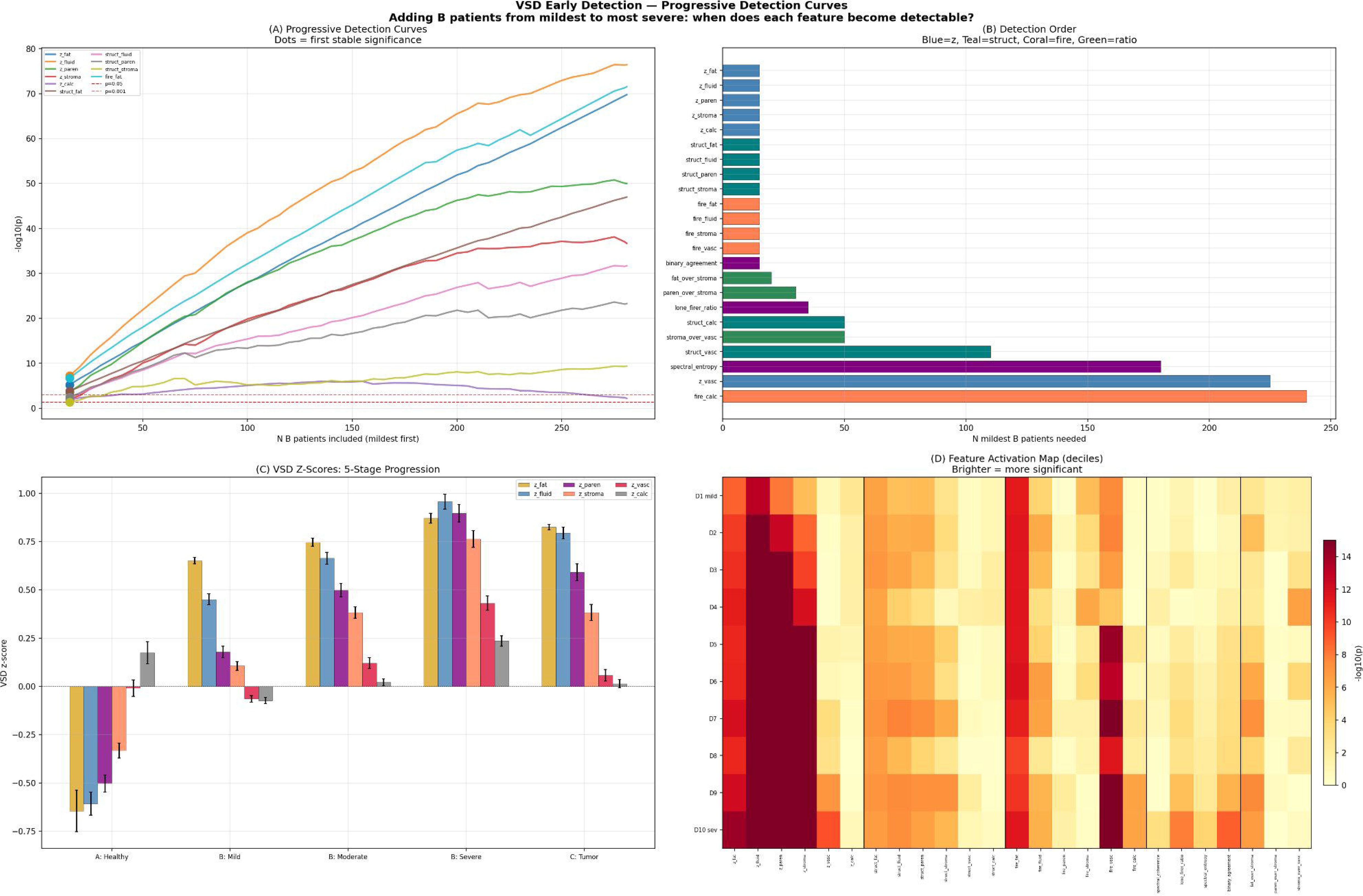
Progressive Detection Curves. (A) p-value curves showing z_fat and z_fluid cross significance earliest when adding B patients from mildest to most severe. (B) Detection order by number of mildest B patients needed for each feature to reach significance. (C) Five-stage VSD z-score progression from healthy through mild, moderate, and severe field effect to tumor. (D) Feature activation heatmap across severity deciles.

**Figure 5.**
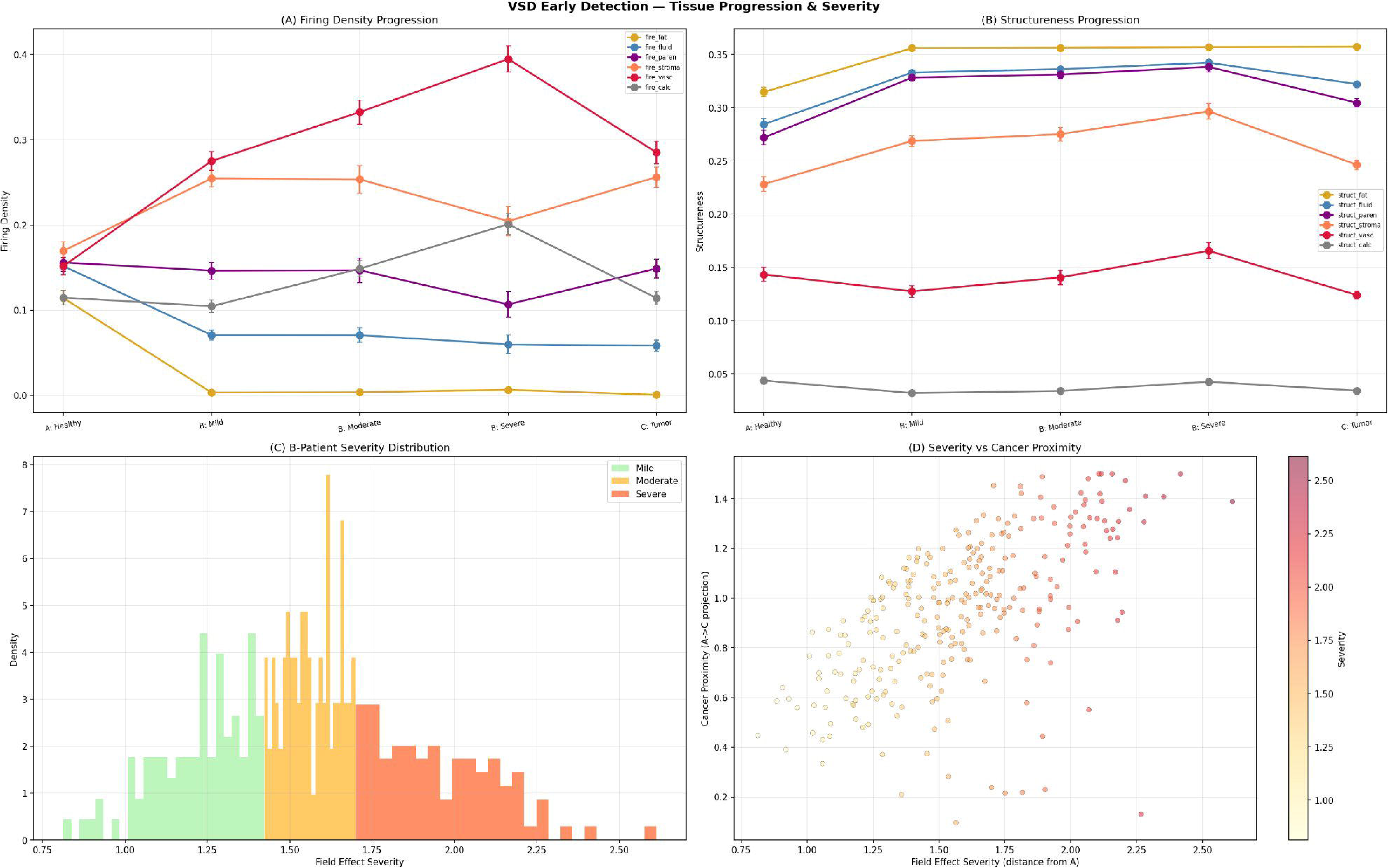
Firing Density and Structureness Progression. (A) Firing density progression across five stages showing fire_stroma and fire_vasc increase while fire_fat collapses. (B) Structureness progression showing structural reorganization. (C) B-patient severity distribution. (D) Severity vs. cancer proximity scatter plot.

### 3.5 Multi-Institutional Validation (CPTAC-PDA)

On the CPTAC-PDA dataset (82 patients, multi-institutional, auto-segmented), VSD achieved AUC 0.961 ± 0.001 using only the 6 VSD z-scores and AUC 0.979 ± 0.001 using all 25 features (Figure 6). The same six sigmoid parameters were used without retraining or domain adaptation. Feature-level comparison confirmed consistent directionality: z_fat (d=−2.10 on CPTAC vs. −2.90 on MSD), z_fluid (d=−2.76 vs. −3.10), fire_fat (d=+2.18 vs. +2.95). z_vasc remained non-significant (d=−0.10, p=0.55), confirming it as a late-stage change consistent with the four-stage model (Table 2).

**Figure 6.**
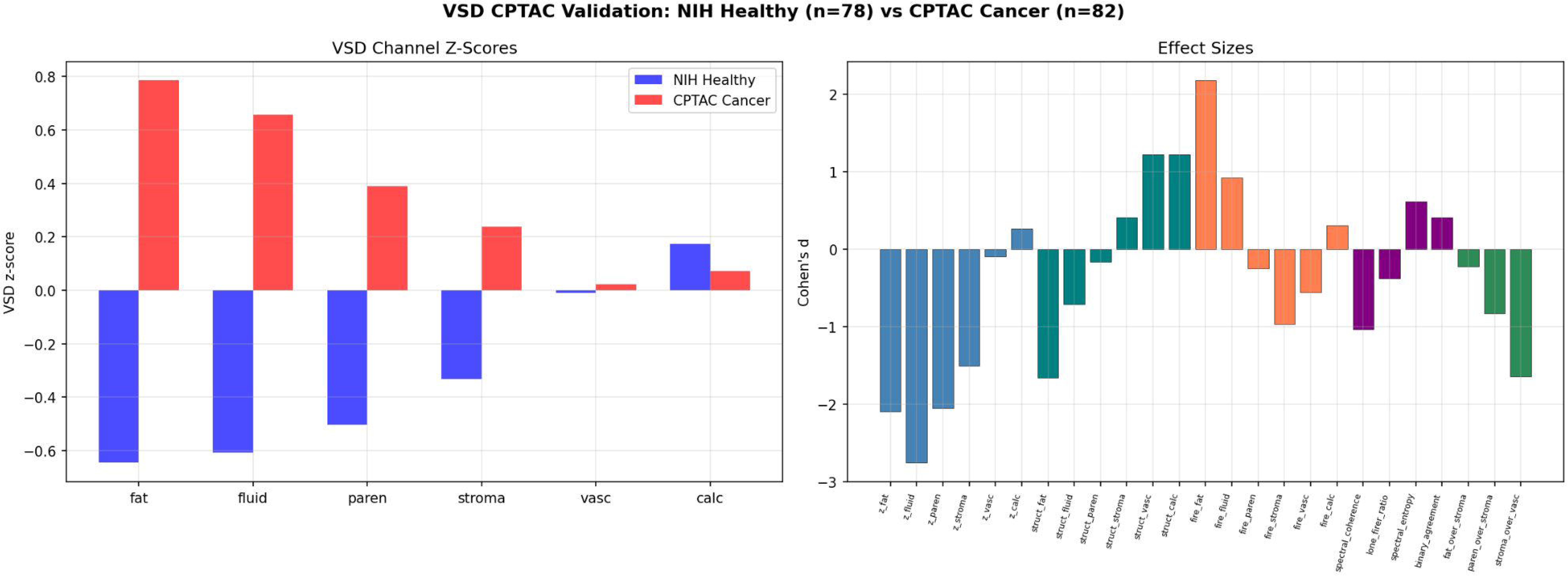
CPTAC Validation. Left: VSD channel z-scores for NIH Healthy (n=78) vs. CPTAC Cancer (n=82) showing the same healthy-vs-cancer pattern as MSD. Right: Cohen’s d effect sizes consistent across all 25 features between datasets.

### 3.6 Temporal Analysis of VSD Signal on CPTAC

CPTAC DICOM time point tags revealed that the 82 analyzed patients were scanned between day −1,394 and day +249 relative to pathological diagnosis, with 74 patients scanned before pathological confirmation. VSD severity showed no correlation with days-from-pathological-diagnosis (Spearman r=−0.008, p=0.944; Figure 7A). No individual VSD feature showed significant temporal correlation (all p>0.14).

**Figure 7.**
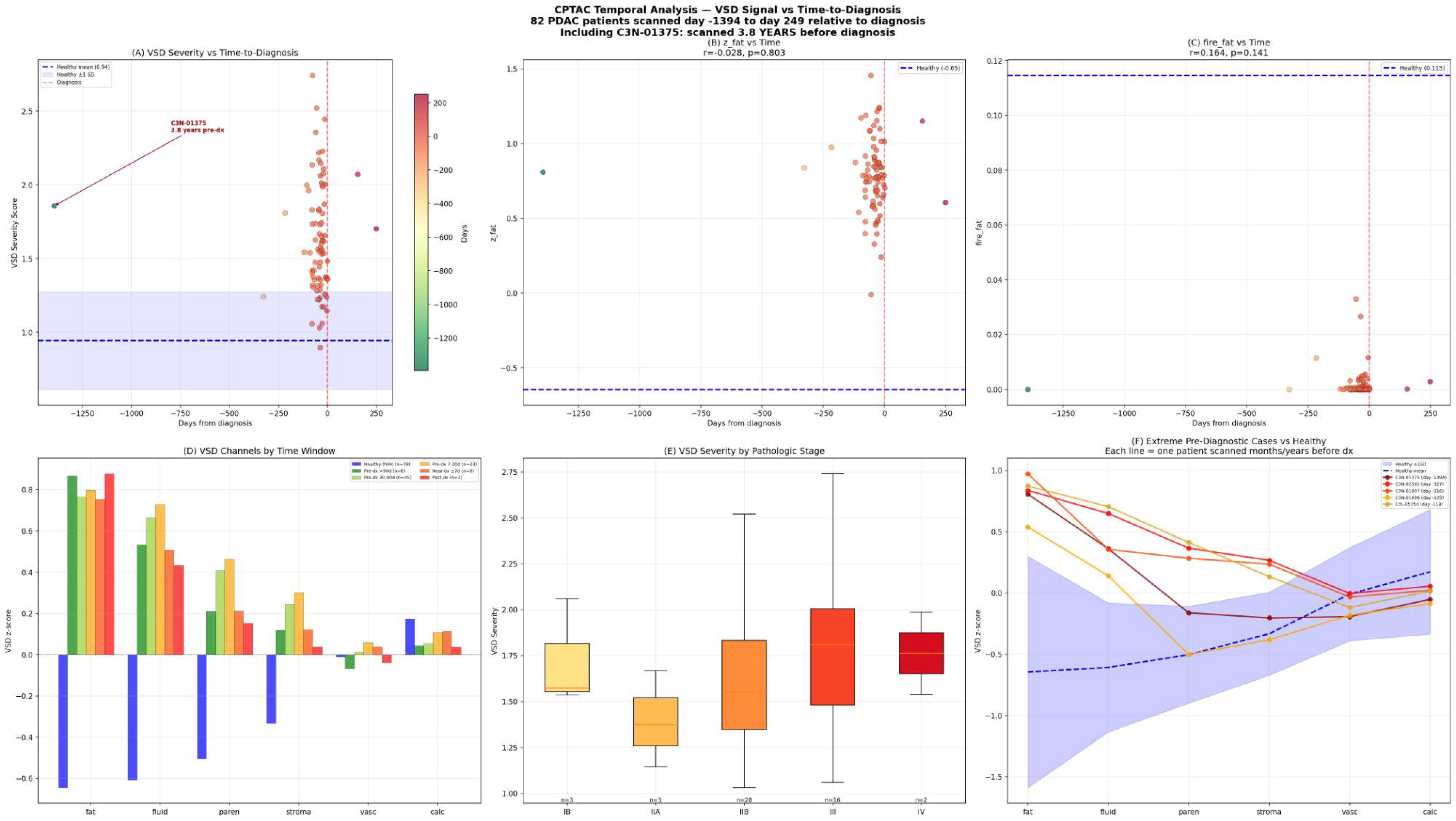
CPTAC Temporal Analysis. (A) VSD severity vs. days-from-pathological-diagnosis—flat curve (r=−0.008, p=0.944), with C3N-01375 (3.8 years pre-dx) annotated. Note: most scans are staging CTs obtained after clinical suspicion (see Discussion 4.2). (B) z_fat vs. time. (C) fire_fat vs. time. (D) VSD channels by time window showing no significant differences between early and late pre-pathological subgroups. (E) Severity by pathologic stage. (F) Extreme pre-diagnostic cases plotted against the healthy reference band.

In an exploratory observation, patient C3N-01375 was scanned 1,394 days (3.8 years) before pathological diagnosis with VSD severity 1.87, compared to the healthy mean of 0.94±0.33 (z-score from healthy: 2.8). However, **this represents a single case, and without clinical notes we cannot confirm whether this scan was a truly incidental CT or part of an early diagnostic workup for a suspected lesion.** The clinical status of this patient at the time of scanning is unknown.

Comparison of patients scanned >90 days before diagnosis (n=8) with patients scanned within 7 days of diagnosis (n=10) showed no significant differences in VSD profiles (Figure 7D). While this is consistent with temporal stability, **the small subgroup sizes (n=8 vs. n=10) provide limited statistical power to detect temporal trends, and the majority of these scans were likely obtained after clinical suspicion was raised.** This analysis should be interpreted as hypothesis-generating rather than confirmatory.

### 3.7 Clinical Correlations

Merging CPTAC VSD profiles with clinical data from the Proteomic Data Commons revealed that VSD severity correlated with pathologic stage (Figure 7E): Stage IB patients showed lower severity (∼1.45) than Stage III/IV patients (∼1.80). VSD severity did not significantly differ between patients who survived and those who died, nor between those who recurred and those who did not, suggesting that VSD measures the tissue transformation state rather than individual prognosis.

## 4. DISCUSSION

We present Virtual Spectral Decomposition with Dendritic Binary Gating, a computational framework that transforms standard single-energy CT into a six-channel tissue-composition analysis achieving AUC 0.961 for detecting pancreatic tissue transformation on multi-institutional imaging, validated across three independent datasets without parameter retraining. The key innovation is the combination of high classification accuracy with full tissue-level interpretability through a transparent sigmoid-to-tissue mapping chain.

### 4.1 VSD as Computational Histology

VSD occupies a fundamentally different position in the radiomics landscape from both conventional texture analysis and deep learning. Conventional radiomics (GLCM, GLRLM, wavelet features) operates on raw grayscale images, producing mathematical descriptors without tissue-specific meaning. Deep learning operates on raw pixels, learning representations that are effective but opaque. VSD operates on the HU spectral axis, producing channels that correspond to named tissue components with known histological correlates: fat (μ=−60) maps to interlobular adipose tissue, stroma (μ=75) maps to collagen-rich desmoplasia, vascular (μ=130) maps to contrast-enhanced vasculature. This is not learned representation—it is encoded domain knowledge from established radiology physics.

The analogy to optical spectroscopy is precise. In spectrophotometry, one decomposes broadband absorption into wavelength-specific channels to identify chromophore concentrations. In VSD, one decomposes the HU spectrum into tissue-specific channels to identify tissue-component concentrations. The sigmoid function serves the same role as a bandpass filter: it isolates the response of interest while suppressing contributions from neighboring tissue types.

The existing patent application (18/895,105) describes this as a generalization of the CIELAB color space transformation used in fundus photography—CIELAB uses 3 fixed perceptual channels; VSD uses N tunable tissue-specific sigmoid channels.

### 4.2 Temporal Observations and Their Limitations

The CPTAC temporal analysis yielded an intriguing but preliminary finding: VSD severity was not correlated with time-to-pathological-diagnosis across the available range (r=−0.008, p=0.944). **However, several important caveats constrain the interpretation of this result.**

First, the CPTAC time point tags measure days before *pathological confirmation*, not days before cancer was first clinically suspected. The majority of scans in this dataset are staging CTs obtained after clinical suspicion was raised—meaning the cancer was almost certainly present at the time of scanning. This is fundamentally different from detecting cancer on truly incidental pre-diagnostic imaging, which is the clinical use case of greatest interest.

Second, the subgroup analysis comparing early (>90 days pre-diagnosis, n=8) and late (≤7 days pre-diagnosis, n=10) time windows has very limited statistical power. The absence of a significant difference between these groups cannot be interpreted as evidence of temporal stability; the sample sizes are insufficient to rule out clinically meaningful differences.

Third, the observation that patient C3N-01375 showed elevated VSD severity 3.8 years before pathological confirmation is noteworthy but represents a single case. Without access to clinical notes, we cannot determine whether this scan was a truly incidental CT (the scenario relevant to screening) or an early workup for a suspected pancreatic lesion. This case should be regarded as hypothesis-generating.

Taken together, the temporal data are consistent with the hypothesis that VSD detects an early, persistent tissue state—potentially corresponding to the PanIN-2/3 microenvironment remodeling that is known to precede invasive carcinoma by years. However, **this hypothesis requires prospective validation on a cohort of patients with truly incidental pre-diagnostic CTs (e.g., from new-onset diabetes surveillance or IPMN monitoring programs) to be confirmed.** We explicitly refrain from claiming a validated detection window based on the current data.

### 4.3 Comparison to Existing Methods

*Pre-diagnostic AI approaches.* Qureshi et al. (Cedars-Sinai) achieved 86% accuracy on pre-diagnostic CTs up to 3 years before diagnosis using radiomic texture features in 36 patients [9]. Kenner et al. (REDMOD) achieved AUC 0.98 using 88 features [8]. VSD achieves comparable discrimination (AUC 0.961) with dramatically fewer features (6–25) that are individually interpretable in tissue-specific terms. Unlike these approaches, VSD’s features have direct histological correlates, enabling clinical reasoning about what tissue changes are being detected.

*Large-scale detection approaches.* Cao et al. (PANDA) achieved AUC 0.986–0.996 for lesion detection on non-contrast CT across 6,239 patients and has received FDA Breakthrough Device Designation [7]. PANDA is a detection tool for existing lesions; VSD is designed to detect the field effect of tissue transformation that may precede a visible lesion. These are complementary rather than competing approaches, and VSD’s pre-lesion detection hypothesis awaits prospective validation.

*Retrospective radiological studies.* Ahmed et al. reported that human radiologists could retrospectively identify pancreatic abnormalities on CT in up to 38% of cases 5 years before PDAC diagnosis [10], and Toshima et al. found abnormalities in 65% of scans 1 year before Stage 1 PDAC [11]. These studies establish that pre-diagnostic CT changes exist but rely on expert retrospective review. VSD offers an automated, quantitative approach to detecting such changes.

The Dendritic Binary Gating component adds a capability absent from all prior radiomics methods: per-channel structural analysis and cross-channel architectural coordination. The co-firing analysis and lone firer signature have no analogues in conventional radiomics, providing the architectural dimension of tissue assessment—not just what tissue components are present, but how they are spatially organized.

### 4.4 Limitations

This study has several important limitations. First, all analyses are retrospective. Second, and most critically, CPTAC scans are predominantly staging CTs rather than truly pre-cancer incidental CTs; the temporal analysis therefore demonstrates that VSD detects tissue changes in patients with known (but not yet pathologically confirmed) PDAC, not that VSD can detect changes before cancer is clinically suspected. Prospective validation on incidental CT cohorts (new-onset diabetes surveillance, IPMN monitoring, routine abdominal CT for other indications) is essential to establish the pre-cancer detection capability. Third, the CPTAC temporal subgroups are small (n=8 and n=10), providing insufficient power for definitive temporal analysis. Fourth, CPTAC auto-segmentation introduces noise compared to manual expert segmentation. Fifth, the study analyzes portal-venous phase CT; performance on arterial, delayed, or non-contrast phases has not been validated. Sixth, the sigmoid parameters were selected based on published HU ranges rather than optimized; data-driven parameter optimization might improve performance but would reduce the physics-based interpretability advantage.

### 4.5 Future Directions and Partnership Opportunities

The critical next step is prospective longitudinal validation. Ideal validation cohorts would include: (a) patients undergoing routine abdominal CT for non-pancreatic indications who subsequently develop PDAC, providing truly incidental pre-diagnostic imaging; (b) high-risk surveillance cohorts (familial pancreatic cancer, CDKN2A carriers, new-onset diabetes) undergoing serial imaging; and (c) IPMN monitoring cohorts where serial CT is standard of care. Several existing resources could support this validation, including the PRECEDE Consortium, institutional longitudinal CT databases at high-volume pancreatic cancer centers, and the Pancreatic Cancer Detection Consortium (PCDC) prospective cohorts.

The VSD framework is modality-agnostic: the same sigmoid decomposition applies to any scalar measurement axis where different tissue components occupy different ranges. Application to other CT-imaged cancers (lung, liver, renal) requires only organ-specific parameter selection.

Additionally, correlation of VSD channel responses with CPTAC proteomic data could provide direct molecular validation of the tissue-component interpretations. Finally, VSD-decomposed multi-channel images could serve as interpretable inputs to spatial deep learning architectures, combining VSD’s tissue-specific decomposition with CNN’s spatial pattern recognition.

## 5. CONCLUSIONS

Virtual Spectral Decomposition with Dendritic Binary Gating detects pancreatic cancer tissue composition changes with AUC 0.961 using six interpretable sigmoid features on standard CT, validated across three independent datasets (NIH, MSD, CPTAC-PDA) without parameter retraining. The VSD tissue transformation signature was temporally stable across available pre-pathological time points in the CPTAC cohort, though the majority of these scans were obtained after clinical suspicion was raised. An exploratory observation of elevated VSD severity 3.8 years before pathological confirmation in one patient, combined with established PanIN biology, suggests the hypothesis of a multi-year pre-diagnostic detection window—but this remains unvalidated. VSD provides a fully transparent sigmoid-to-tissue interpretability chain, requires no special hardware or training data, and produces tissue-specific features with direct histological correlates. Prospective validation on truly incidental pre-diagnostic CT cohorts is warranted and represents the essential next step toward establishing VSD as a screening tool for early pancreatic cancer detection.

## Data Availability

All datasets analyzed are publicly available: NIH Pancreas-CT (Roth et al., 2016), MSD Task07 (Antonelli et al., 2022), and CPTAC-PDA (DOI: 10.7937/K9/TCIA.2018.SC20FO18) from The Cancer Imaging Archive. Code will be made available upon reasonable request.

## Ethics Statement

This study analyzed publicly available, de-identified datasets. No IRB approval was required.

## Data Availability

All source data analyzed in this study are publicly available. CT imaging data: NIH Pancreas-CT (https://wiki.cancerimagingarchive.net/display/Public/Pancreas-CT), Medical Segmentation Decathlon Task07 (http://medicaldecathlon.com/), and CPTAC-PDA (https://doi.org/10.7937/K9/TCIA.2018.SC20FO18) from The Cancer Imaging Archive. Clinical data: Proteomic Data Commons (https://pdc.cancer.gov, Study ID: PDC000270). Derived VSD feature profiles and analysis code are available upon reasonable request to the authors.

https://www.cancerimagingarchive.net/collection/cptac-pda/

http://medicaldecathlon.com/

https://pdc.cancer.gov/pdc/study/PDC000270

## References

[1] Siegel RL, Miller KD, Wagle NS, Jemal A. Cancer statistics, 2023. CA Cancer J Clin. 2023;73(1):17–48.

[2] Rahib L, Smith BD, Aizenberg R, et al. Projecting cancer incidence and deaths to 2030: the unexpected burden of thyroid, liver, and pancreas cancers in the United States. Cancer Res. 2014;74(11):2913–2921.

[3] Yachida S, Jones S, Bozic I, et al. Distant metastasis occurs late during the genetic evolution of pancreatic cancer. Nature. 2010;467(7319):1114–1117.

[4] Roth HR, Lu L, Farag A, et al. DeepOrgan: Multi-level deep convolutional networks for automated pancreas segmentation. Proceedings of MICCAI. 2015.

[5] Antonelli M, Reinke A, Bakas S, et al. The Medical Segmentation Decathlon. Nat Commun. 2022;13(1):4128.

[6] National Cancer Institute Clinical Proteomic Tumor Analysis Consortium (CPTAC). The Clinical Proteomic Tumor Analysis Consortium Pancreatic Ductal Adenocarcinoma Collection (CPTAC-PDA). The Cancer Imaging Archive. DOI: 10.7937/K9/TCIA.2018.SC20FO18.

[7] Cao K, Xia Y, Yao J, et al. Large-scale pancreatic cancer detection via non-contrast CT and deep learning. Nat Med. 2023;29(12):3033–3043.

[8] Kenner B, Chari ST, Kelsen D, et al. Artificial intelligence and early detection of pancreatic cancer. Pancreas. 2021;50(3):251–279.

[9] Qureshi TA, Gaddam S, Wachsman AM, et al. Predicting pancreatic ductal adenocarcinoma using artificial intelligence analysis of pre-diagnostic computed tomography images. Cancer Biomark. 2022;33(2):211–217.

[10] Ahmed TM, Chu LC, Javed AA, et al. Hidden in plain sight: commonly missed early signs of pancreatic cancer on CT. Abdom Radiol. 2024;49(10):3599–3614.

[11] Toshima F, Watanabe R, Inoue D, et al. CT abnormalities of the pancreas associated with the subsequent diagnosis of clinical stage I pancreatic ductal adenocarcinoma more than 1 year later: a case-control study. AJR Am J Roentgenol. 2021;217(6):1353–1364.

[12] Wasserthal J, Breit HC, Meyer MT, et al. TotalSegmentator: Robust segmentation of 104 anatomic structures in CT images. Radiology: AI. 2023;5(5):e230024.

[13] Clark K, Vendt B, Smith K, et al. The Cancer Imaging Archive (TCIA): Maintaining and operating a public information repository. J Digit Imaging. 2013;26(6):1045–1057.

